# Treatment Response to Hydroxychloroquine and Antibiotics for mild to moderate COVID-19: a retrospective cohort study from South Korea

**DOI:** 10.1101/2020.07.04.20146548

**Authors:** Min Ho An, Min Seo Kim, Yu-Kyung Park, Bong-Ok Kim, Seok Ho Kang, Won Jun Kim, Sung Kyu Park, Hea-Woon Park, Wonjong Yang, Joonyoung Jang, Soon-Woo Jang, Tae-Ho Hwang

**Affiliations:** Ajou University, School of Medicine, Suwon, Republic of Korea; Korea University, College of Medicine, Seoul, Republic of Korea; Korea Workers’ Compensation & Welfare Services Daegu Hospital, 515 Hakjeong-ro, Buk-gu, Daegu, Republic of Korea; Director of So Ahn Public Health Center, Wando, Republic of Korea; Director of Cheongsan Public Health Center, Wando, Republic of Korea; Director of Bukha Public Health Center, Jangseong, Republic of Korea; Department of Pharmacology, Pusan National University, School of Medicine, Yangsan, Republic of Korea; Gene and Cell Therapy Research Center for Vessel-associated Diseases, School of Medicine, Pusan National University, Yangsan, Republic of Korea; Department of Urology, Korea University, School of Medicine, Seoul, Republic of Korea; Pusan University, School of Medicine, Yangsan, Republic of Korea

**Keywords:** COVID-19, Hydroxychloroquine, antibiotics, azithromycin, treatment response, retrospective cohort study

## Abstract

**Objectives:** To assess the efficacy of hydroxychloroquine on mild-moderate COVID-19 patients in South Korea.

**Methods:** A retrospective cohort study of the 358 laboratory-confirmed SARS-CoV-2 (COVID-19) patients was conducted. 226 patients met inclusion criteria for analysis. Propensity score matching (PSM) and Cox regression method were utilized to control and adjust for confounding factors. Mild to moderate COVID-19 patients were managed with hydroxychloroquine (HQ) plus antibiotics (n = 31) or conservative treatment (n = 195).

**Results:** Kaplan-Meier curves drawn using propensity score-matched data revealed no differences between the length of time to viral clearance and duration of hospital stay between the two treatment arms (p=0.18, p=0.088). Multivariable Cox regression analysis similarly showed that time to viral clearance(Hazard ratio (HR) 0.97, [95%-confidence interval (CI): 0.57-1.67]) and symptom duration(HR 1.05, [95%-CI: 0.62-1.78]) were not different between groups. No severe adverse event or death was observed in either group.

**Conclusions:** HQ with antibiotics was not associated with better clinical outcomes in terms of time to viral clearance, length of hospital stay, and duration of symptoms compared to conservative treatment alone. Large prospective randomized trials are necessary for definitive conclusions.

## INTRODUCTION

As of June 27^th^ 2020, over 9,500,000 confirmed cases and over 490,000 deaths due to Coronavirus Disease 2019 (COVID-19) were reported by the World Health Organization (WHO)(1). The causative virus of this pandemic, SARS-CoV-2, presents an unprecedented challenge to healthcare systems worldwide, but no definitive treatment protocol exists due to the lack of clear understanding of the pathogenesis of the disease or the nature of its causative virus (2).

Based on a recent study from China, about 80% of COVID-19 patients show non-severe symptoms(3) Thus, it is relevant to a large number of patients to investigate potential drugs that may be effective for patient with non-severe disease. Numerous hospitals in South Korea have experience treating mild to moderate COVID-19 with management methods such as standard supportive care alone, hydroxychloroquine (HQ), lopinavir-ritonavir (Lop/R), etc. These treatments can be further classified based on the use of adjunct antibiotics, which were prescribed depending on the patients’ symptoms and comorbidities. While South Korea has been relatively successful in managing the spread of the pandemic and its epidemiologic control strategies well-known, the pharmacological management and treatment of its infected citizens have not been reported as extensively.

Herein, we present our experience on COVID-19 management with pharmacological therapy. This study aims to compare treatment responses of mild to moderate COVID-19 patients who received HQ with antibiotics or conservative treatment.

## PATIENTS

A retrospective cohort study of 358 patients with laboratory-confirmed SARS-CoV-2 infection hospitalized in Korea Worker’s Compensation & Welfare Service Daegu Hospital was conducted. All patients were diagnosed with COVID-19 by real-time reverse-transcriptase polymerase chain reaction (RT-PCR) according to the WHO protocol(4). Patients were admitted to the hospital from February 28, 2020, to April 28, 2020. Patients who received at least three days of HQ treatment or any duration of standard therapy were included. They were further stratified by severity according to the National Institutes of Health (NIH) COVID-19 guideline (5). Individuals without shortness of breath, dyspnea, or abnormal imaging were categorized as mild COVID-19; individuals who have evidence of lower respiratory disease by clinical assessment or imaging and oxygen saturation (SaO_2_) >93% on room air at sea level were categorized as moderate COVID-19; individuals who have respiratory frequency >30 breaths per minute, SaO_2_ ≤93% on room air at sea level, ratio of arterial partial pressure of oxygen to fraction of inspired oxygen (PaO_2_/FiO_2_) <300, or lung infiltrates >50% were categorized as severe COVID-19 (5).

Of 358 COVID-19 patients, 226 patients remained for full analysis after excluding patients. Exclusions were made for patients who did not adhere to treatment protocols, patients with severe symptoms as they were referred to tertiary hospitals for intensive care at an early stage of the management, patients who were referred from another hospital, and patients who switched treatments from HQ to Lop/R. The ethics committee of Pusan National University Yangsan Hospital approved this study and granted a waiver of informed consent from study participants.

## METHODS

The authors reviewed the electronic medical records of included patients and collected epidemiological, clinical, historical, laboratory, and treatment outcomes data. Patient confidentiality was protected by deidentifying patient information. The electronic data was also stored in a locked, password-protected computer. All but 3 patients were discharged from within the follow-up period up to April 28, 2020.

### Procedure

To identify SARS-CoV-2 infection, nasal swab samples were obtained from all patients on admission. Collected swab samples were tested for SARS-CoV-2 using RT-PCR, and complete viral clearance was affirmed by two consecutive negatives on RT-PCR, which was defined by cycle threshold (Ct) value ≥40. Additionally, patients received routine blood and biochemical tests; and for those with radiologic bronchiolitis/pneumonia findings, chest x-rays (CXR) or computed tomography (CT) were taken on a regular basis until lesions were resolved. All CXR and CT images were reviewed by experienced radiologists. The highest level of oxygen supports each patient received during their hospitalization was also recorded. Fever was recorded if a patient’s body temperature arose to 37.5 °C or higher, and information regarding all other COVID-19-related symptoms (cough, chill, myalgia, sputum, dyspnea, nasal discharge, and sore throat) was collected daily through a telephone survey using pre-specified questionnaires. All baseline characteristics were measured at the time of admission in hospital.

HQ was administered when patients were suspected to have pneumonitis or bronchiolitis on CXR or CT. Few patients were initiated on HQ but were shortly switched to Lop/R due to side effects such as nausea or progression of pneumonia; these patients were excluded from our study.

Patients who were given HQ received 200mg HQ tablets twice daily. Azithromycin, when prescribed, was used for up to 5 days and given as 500mg tablets once daily; most patient received azithromycin for 3 days, and only two patients received for 4-5 days of azithromycin. Cefixime, when prescribed, was used until remission of pneumonia and was administered as 100mg tablets twice daily.

### Outcomes

As our target patient population was mild-to-moderate COVID-19 patients, our primary endpoint was the duration of viral clearance (i.e. time from admission to two consecutive negative results on PCR, signified by Ct value ≥ 40); and our secondary endpoints were length of hospital stay and symptom duration (i.e. time from earliest date to last date of any symptoms).

### Statistical Analysis

Continuous variables were reported as mean (standard deviation [SD]), and categorial variables were reported as number (%). Categorical data were compared using the χ2 test or Fisher’s exact test, and continuous variables were analyzed using Student’s t-test or Mann-Whitney U test. Kaplan-Meier curves were generated for primary and secondary endpoints and were compared using the log-rank test. Cox proportional hazard ratio (HR) models were used to determine HRs and 95% confidence intervals (CIs). All tests were 2-sided, and a *P* value less than 0.05 was considered statistically significant. All analyses were conducted using IBM SPSS, version 21.0 (SPSS Inc), or R software, version 3.6.0 (R Foundation for Statistical Computing).

### Propensity Score matching and Cox proportional hazards regression models

Propensity score matching was performed to balance the baseline characteristics of two groups of patients who received HQ or standard therapy. Matched variables include age, sex, severity, WBC, initial lymphocyte count, albumin, and CRP. Patients in the two groups were matched at a 1:1 ratio based on their closest propensity score within a threshold of 0.25. Cox proportional-hazards regression models were used to evaluate the association between hydroxychloroquine use and time to viral clearance and symptom duration. Our multivariable Cox regression model included age, sex, severity, WBC, initial lymphocyte count, albumin, CRP, and duration of Lop/R use. In addition, we utilized propensity-score methods to adjust for the effects of confounding. The individual propensities for administration of HQ were estimated with the use of a multivariable logistic-regression model that included the same covariates as the Cox regression model except duration of Lop/R use owing to the lack of Lop/R treatment in the supportive therapy group.

### Data availability

The research data that support the findings of this study are available from corresponding author upon reasonable request.

## RESULTS

### Baseline demographics and initial laboratory indices of patients

A total of 226 patients were included in this study (**Table 1**) and the enrollment of the study cohort is described in **Figure 1**. Of the 226 patients, 31(13.7%) received HQ and 195(86.3%) did not. The mean age was 35.26 years (standard deviation (SD) 14.22) and 43.48(SD 15.50) in the standard supportive therapy group and the HQ group, respectively. 117(60.0%) were female in standard supportive therapy group, and 26 (83.9%) were female in the HQ group. After propensity score matching, most co-variables did not show significant differences except systolic BP and hematocrit. A total of forty-three patients had comorbidities, including hypertension (n=22, [0.1%]), diabetes (n=6, [0.03%]), dyslipidemia (n=7, [0.03%]), and thyroid disease. (n=8, [0.04%] before matching. The length of time from confirmation of diagnosis to admission was significantly different between standard supportive therapy (4.51 days [SD 3.35]) and HQ group (2.35 days [SD 1.52]) before matching, but the difference disappeared after matching. Average duration from diagnosis to HQ treatment initiation before and after matching were (6.19 days [SD 4.01]) and (6.70, [SD 3.92]) respectively. The distribution of the estimated propensity scores for administration of HQ and standard supportive therapy are shown in **Figure S1**.

**Table 1.** Demographic characteristics, initial laboratory indices – before and after propensity score matching.

|  | Before matching |  |  | After Matching |  |  |
| --- | --- | --- | --- | --- | --- | --- |
|  | Standard | HQ | p-value | Standard | HQ | p-value |
| Number of patients | 195 | 31 |  | 20 | 20 |  |
| Sex, female (%) | 117 (60.0) | 26 (83.9) | 0.010 | 19 (95.0) | 16 (80.0) | 0.342 |
| Age (mean (SD)) | 35.26 (14.22) | 43.48 (15.50) | 0.003 | 38.65 (14.88) | 38.50 (16.45) | 0.976 |
| BMI | 22.95 (3.20) | 23.24 (3.24) | 0.652 | 22.61 (3.31) | 23.32 (3.67) | 0.540 |
| Fever >= 37.5 (%) | 24 (12.3) | 9 (29.0) | 0.025 | 5 (25.0) | 4 (20.0) | 1.000 |
| Asymptomatic (%) | 37 (19.0) | 1 (3.2) | 0.029 | 0 (0.0) | 1 (5.0) | 1.000 |
| O2 saturation =< 95 (%) | 14 (7.2) | 2 (6.5) | 1.000 | 5 (25.0) | 2 (10.0) | 0.407 |
| O2 supply application (%) | 0 (0.0) | 1 (3.2) | 0.137 | 0 (0.0) | 0(0.0) | 1.000 |
| Moderate severity (%) | 46 (23.6) | 31 (100.0) | <0.001 | 20 (100.0) | 20 (100.0) | 1.000 |
| **Duration of treatment delay** |  |  |  |  |  |  |
| Confirmatory diagnosis to HQ treatment, days | 0.00 (0.00) | 6.19(4.01) | <0.001 | 0.00 (0.00) | 6.70(3.92) | <0.001 |
| Confirmatory diagnosis to admission, days | 4.51(4.35) | 2.35(1.52) | <0.001 | 4.10(4.41) | 2.60(1.64) | 0.162 |
| **Comorbidities** |  |  |  |  |  |  |
| Hypertension (%) | 20 (10.3) | 2 (6.5) | 0.747 | 2 (10.0) | 2 (10.0) | 1.000 |
| Dyslipidemia (%) | 6 (3.1) | 1 (3.2) | 1.000 | 0 (0.0) | 0 (0.0) | 1.000 |
| Diabetes mellitus (%) | 6 (3.1) | 0 (0.0) | 1.000 | 0 (0.0) | 0 (0.0) | 1.000 |
| Thyroid (%) | 6 (3.1) | 2 (6.5) | 0.302 | 1 (5.0) | 1 (5.0) | 1.000 |
| **Vital Signs** |  |  |  |  |  |  |
| Systolic BP (mean (SD)) | 128.54 (16.34) | 129.35 (17.91) | 0.799 | 115.00 (26.03) | 129.30 (16.98) | 0.047 |
| Diastolic BP (mean (SD)) | 77.73 (10.41) | 79.81 (15.33) | 0.472 | 72.70 (7.51) | 77.85 (15.84) | 0.197 |
| Heart rate (mean (SD)) | 86.86 (11.91) | 87.52 (12.67) | 0.777 | 86.95 (9.86) | 85.05 (13.76) | 0.619 |
| Respiratory rate (mean (SD)) | 20.06 (0.71) | 20.13 (0.50) | 0.585 | 19.80 (0.89) | 20.10 (0.45) | 0.188 |
| Body temperature (mean (SD)) | 36.99 (0.40) | 37.25 (0.57) | 0.019 | 37.06 (0.47) | 37.08 (0.44) | 0.890 |
| **Laboratory indices** |  |  |  |  |  |  |
| White blood cells (mean (SD)) | 6.05 (1.56) | 5.28 (1.52) | 0.012 | 5.42 (1.00) | 5.73 (1.49) | 0.436 |
| Lymphocytes (mean (SD)) | 2.03 (0.52) | 1.73 (0.56) | 0.003 | 1.85 (0.46) | 1.87 (0.59) | 0.946 |
| Red blood cells (mean (SD)) | 4.70 (0.52) | 4.52 (0.52) | 0.071 | 4.29 (0.35) | 4.65 (0.54) | 0.016 |
| Hemoglobin (mean (SD)) | 14.07 (1.68) | 13.48 (1.59) | 0.067 | 13.14 (1.06) | 13.90 (1.73) | 0.099 |
| Hematocrit (mean (SD)) | 42.26 (4.43) | 40.78 (4.31) | 0.085 | 39.29 (2.78) | 41.91 (4.58) | 0.035 |
| Cr (mean (SD)) | 0.79 (0.18) | 0.74 (0.17) | 0.186 | 0.68 (0.11) | 0.76 (0.19) | 0.096 |
| BUN (mean (SD)) | 12.24 (3.03) | 11.67 (3.36) | 0.338 | 11.73 (3.78) | 11.19 (2.85) | 0.606 |
| AST (mean (SD)) | 25.10 (21.06) | 26.19 (15.50) | 0.781 | 19.75 (7.64) | 28.05 (18.42) | 0.074 |
| ALT (mean (SD)) | 26.30 (22.79) | 30.77 (45.76) | 0.597 | 19.15 (11.94) | 38.55 (55.60) | 0.142 |
| Total bilirubin (mean (SD)) | 0.56 (0.30) | 0.57 (0.52) | 0.961 | 0.45 (0.31) | 0.60 (0.61) | 0.361 |
| Albumin (mean (SD)) | 4.36 (0.28) | 4.07 (0.32) | <0.001 | 4.15 (0.19) | 4.14 (0.33) | 0.968 |
| Platelet (mean (SD)) | 266.66 (62.35) | 259.35 (63.16) | 0.546 | 269.85 (74.69) | 256.85 (66.80) | 0.565 |
| LDH (mean (SD)) | 221.60 (79.16) | 263.65 (118.39) | 0.064 | 198.85 (40.37) | 254.15 (136.17) | 0.095 |
| PT(INR) (mean (SD)) | 1.02 (0.06) | 1.03 (0.06) | 0.434 | 1.04 (0.08) | 1.04 (0.07) | 0.883 |
| Total cholesterol (mean (SD)) | 161.94 (32.91) | 155.87 (27.47) | 0.331 | 152.65 (26.65) | 157.05 (24.59) | 0.591 |
| HDL (mean (SD)) | 45.87 (11.25) | 44.00 (8.83) | 0.378 | 45.13 (9.23) | 43.76 (8.22) | 0.622 |
| TG (mean (SD)) | 164.78 (75.29) | 151.45 (50.06) | 0.342 | 153.45 (105.30) | 151.95 (55.17) | 0.955 |
| Glucose (mean (SD)) | 94.36 (45.87) | 99.74 (34.38) | 0.533 | 83.50 (31.10) | 100.55 (40.12) | 0.141 |
| CRP (mean (SD)) | 0.17 (0.31) | 0.89 (1.77) | 0.030 | 0.17 (0.20) | 0.61 (1.86) | 0.300 |

BMI: body mass index; HQ: hydroxychloroquine; AST: Aspartate aminotransferase; ALT: alanine aminotransferase; LDH: lactate dehydrogenase; BUN: blood urea nitrogen; HDL: high-density lipoproteins; PT (INR): prothrombin time (international normalized ratio); CRP: c-reactive protein; TG: triacyl-glyceride; SD: standard deviation.

**Figure 1.**
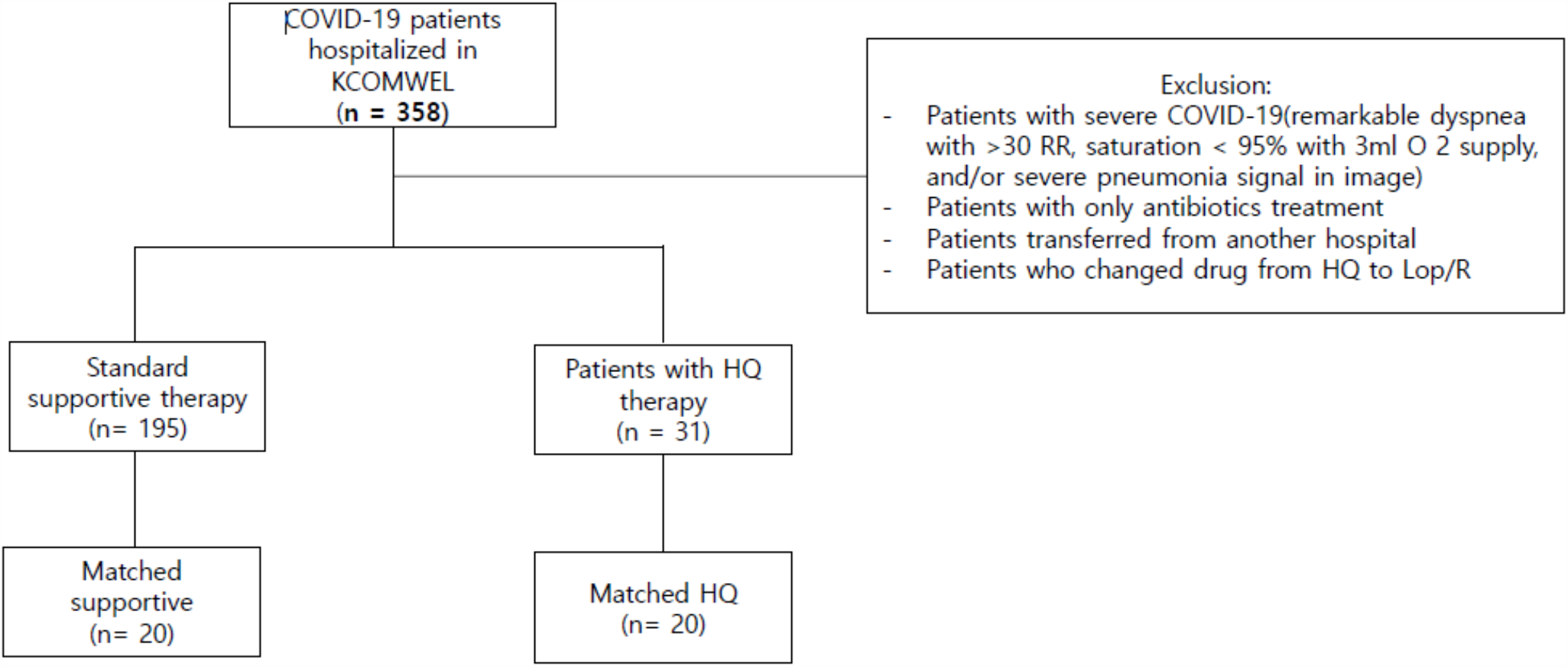
Flowchart for enrollment of the study cohort.

### General clinical outcomes and adverse reactions

There were significant differences between two groups before propensity score matching in all endpoints: time to viral clearance, hospital stay, and symptom duration (**Table 2**). However, these differences faded after propensity score matching. No significant difference in adverse reactions were observed in both groups before and after matching. Most frequently observed side effect was increased AST/ALT in both groups.

**Table 2.** Clinical outcomes, adverse reactions, and duration of medication use before and after propensity score matching.

|  | Before matching |  |  | After Matching |  |  |
| --- | --- | --- | --- | --- | --- | --- |
|  | Standard | HQ | p-value | Standard | HQ | p-value |
| Number of patients | 195 | 31 |  | 20 | 20 |  |
| **Clinical outcomes** |  |  |  |  |  |  |
| Mortality (%) | 0 (0) | 0 (0) | 1.000 | 0 (0) | 0 (0) | 1.000 |
| Viral clearance duration<br>(mean (SD)), days | 18.82 (6.52) | 22.68 (5.42) | 0.002 | 21.35 (4.69) | 23.45 (5.68) | 0.211 |
| Hospital stay (mean (SD)),<br>days | 15.43 (6.24) | 21.52 (5.34) | <0.001 | 19.45 (4.43) | 22.10 (5.51) | 0.102 |
| Symptom resolution duration<br>(mean (SD)), days | 11.45 (9.67) | 18.00 (8.29) | <0.001 | 18.95 (6.96) | 14.55 (8.08) | 0.073 |
| **Adverse reactions** |  |  |  |  |  |  |
| Nausea and Vomiting (%) | 0 (0.0) | 1 (3.2) | 0.137 | 0 (0.0) | 1 (5.0) | 1.000 |
| Abdominal discomfort<br>/diarrhea (%) | 0 (0.0) | 1 (3.2) | 0.137 | 0 (0.0) | 1 (5.0) | 1.000 |
| Increased total bilirubin (%) | 0 (0.0) | 1 (3.2) | 0.137 | 0 (0.0) | 1 (5.0) | 1.000 |
| Increased BUN (%) | 0 (0.0) | 0 (0.0) | 1.000 | 0 (0.0) | 0 (0.0) | 1.000 |
| Increased AST/ALT (%) | 1 (0.5) | 4 (12.9) | 0.001 | 1 (5.0) | 3 (15.0) | 0.605 |
| **Duration of medication use** |  |  |  |  |  |  |
| Cefixime use (mean (SD)), days | 0.00 (0.00) | 10.90 (4.26) | <0.001 | 0.00 (0.00) | 10.10 (3.80) | <0.001 |
| AZ use (mean (SD)), days | 0.00 (0.00) | 3.03 (0.55) | <0.001 | 0.00 (0.00) | 3.05 (0.69) | <0.001 |
| HQ use (mean (SD)), days | 0.00 (0.00) | 8.52 (3.09) | <0.001 | 0.00 (0.00) | 8.85 (3.10) | <0.001 |

HQ: hydroxychloroquine; Lop/R: lopinavir-ritonavir; AST: Aspartate aminotransferase; ALT: alanine aminotransferase; BUN: blood urea nitrogen; SD: standard deviation.

### Treatment response

Although the significant differences were observed in length of time to viral clearance (log -rank p =0.02) and hospital stay (log -rank p < 0.001) in the Kaplan-Meier curves before propensity score matching, the differences were no longer observed after patients were matched for propensity scores (**Figure 2**).

**Figure 2.**
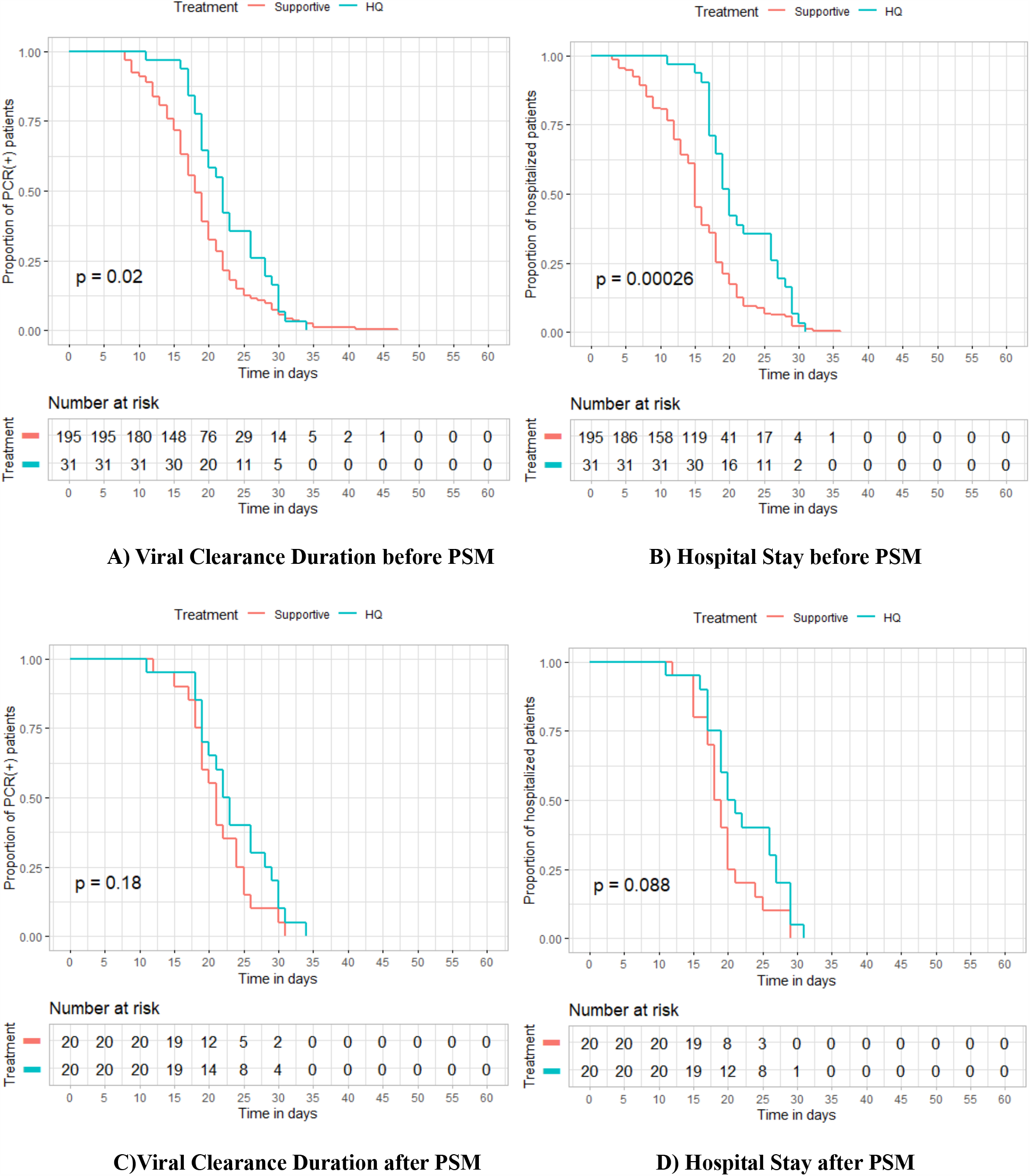
Kaplan-Meier curves for viral clearance duration and hospital stay before and after propensity score matching.

In the crude unadjusted analysis, the length of time to viral clearance in patients who had received HQ were shown to be less likely to achieve viral clearance (Hazard ratio (HR), 1.53[95%-CI: 1.04-2.22]) and symptom resolution (HR, 1.61[95%-CI: 1.1-2.32]) compared to standard supportive therapy (**Table 3**). However, no significant difference was observed in time to viral clearance (HR, 0.97[95%-CI: 0.57-1.67]) and in symptom resolution (HR, 1.05[95%-CI: 0.62-1.78]) in multivariable analysis. Additionally, cox regression analysis was performed with calculated propensity score as an adjust variable, and no significant differences were observed for both outcomes. Further analysis was performed with matched population, and no significant differences were observed in the adjusted analysis (**Table 3**).

**Table 3.** Associations between hydroxychloroquine use and time to viral clearance and symptom duration in crude analysis, multivariable analysis, and propensity-score matching compare to standard supportive therapy. (Conservative therapy is the reference)

|  | **Hazard ratio** | **95% CI** | **p** |
| --- | --- | --- | --- |
| **Time to viral clearance** |  |  |  |
| Univariable Cox regression analysis (n=31) | 1.53 | 1.04-2.22 | 0.028 |
| Multivariate Cox regression analysis (n=31) * | 0.97 | 0.57-1.67 | 0.918 |
| Multivariate Cox regression analysis with PSM (n=31) | 1.16 | 0.67-2 | 0.569 |
| Cox regression with matched population (n=20) ** | 1.53 | 0.83-2.94 | 0.184 |
| **Time to Symptom resolution** |  |  |  |
| Univariable Cox regression analysis (n=31) | 1.61 | 1.1-2.32 | 0.015 |
| Multivariate Cox regression analysis (n=31) * | 1.05 | 0.62-1.78 | 0.859 |
| Multivariate Cox regression analysis adjusted with PSM (n=31) | 0.85 | 0.48-1.5 | 0.589 |
| Cox regression with matched population (n=20) ** | 1.09 | 0.59 - 2.08 | 0.79 |

\* adjusted for age, sex, moderate severity, WBC, initial lymphocyte count, albumin, CRP, duration of Lop/R use; \*\*matched with age, sex, moderate severity, WBC, initial lymphocyte count, albumin, CRP; PSM: propensity score matching

## DISCUSSION

The present study represents the largest pharmacological study on the management of COVID-19 from South Korea. This retrospective cohort study compared treatment response to two different treatment protocols in mild to moderate COVID-19 patients using several clinical outcome measures. HQ plus antibiotics did not show clinical improvement compared to conservative treatment in terms of viral clearance, hospital stay, and symptom duration.

It is notable that HQ plus antibiotics group had worse baseline clinical profiles (i.e. higher percentage of moderate severity patients, more patients with fever >=37.5C, higher average body temperature) and prognostic indicators such as age, LDH, lymphocyte count, and CRP (**Table 1**)(6). However, we have attempted to account for this difference using propensity score matching, and patient characteristics were balanced after matching with propensity scores. Other confounder-adjusting analyses were performed as well (**Table 3**), and the results consistently reported no significant difference in the clinical outcomes between HQ and standard care groups.

HQ became the primary treatment option at the beginning of the pandemic as it was shown to be effective in several in-vitro studies against SARS-CoV-2(7-9) and a non-randomized trial conducted by Gautret et al showing superior viral clearance in group treated with HQ(10). Moreover, a few trials have shown significant reduction in viral-load (10), earlier time to symptom resolution(11), and improvement in chest radiographs (11). However, recent studies conducted with more controlled designs and larger sample sizes have produced conflicting results: a recent study by Tang et al. reported no differences in negative conversion rate in patients treated with HQ compared to standard supportive treatment (12); study by Geleris et al that analyzed 1376 patients concluded no beneficial effect of HQ on patients’ composite outcome of mortality and progression to severe disease(13); and an observational comparative study on patients with COVID-19 pneumonia revealed that administration of HQ was not associated with reduction in mortality or intensive care unit admissions (14). Our result is in accordance with these latest studies. However, more data must be accrued to draw definitive conclusions, and clinicians must be mindful that the evidence behind treatment of COVID-19 is still incomplete and under investigation.

Azithromycin and/or cefixime were prescribed in addition to HQ in our study cohort for management of pneumonia and bacterial co-infection as per recommendations from the Korean Society of Infectious Disease(15) and other literature (16-20). Combination of HQ and azithromycin has gained attention for its potential synergistic therapeutic effect on managing COVID-19 as both drugs were proposed to act as competitive inhibitors of SARS-CoV-2 attachment to the host-cell membrane (21); on the other hand, the potential cardiotoxicity after taking HQ and azithromycin raised concern from many clinicians as multiple studies presented accumulating evidence of higher risk for serious adverse events such as torsades de pointes and ventricular arrhythmia in those who were prescribed the combination therapy (22-24). Our study did not include heart rhythm monitoring for patients and is thus unable to add to the safety aspect of the discussion.

In our cohort, no mortality or serious adverse effects were observed. A total of 8 cases of minor adverse reactions to treatment were reported (8 out of 226 cases). The most commonly reported adverse reaction was increased AST/ALT. However, all patients with adverse reactions were discharged without any harmful sequelae. AST/ALT returned to normal before discharge in all patients except one, but this patient had elevated AST/ALT at baseline. The absence of serious adverse events may be attributable to our protocol, which prescribed reduced dosages of medications administered at once (i.e. 200mg HQ tablet per each) by using a twice daily regimen.

While increasing evidence claims that HQ is limited in its efficacy as a treatment of COVID-19, its use is still being investigated for prevention. The first randomized controlled trial investigating prophylactic HQ conducted by Boulware et al revealed that there was no preventive effect in taking 600mg HQ within 4 days after of exposure; it is also noteworthy that 40% of patients who received a prophylactic dose experienced side effects such as nausea, vomiting, and diarrhea, while only 16% experienced adverse events in the control group(25). Numerous trials on prophylactic HQ are underway (26), and future studies should enable more conclusive statements on the prophylactic effect of HQ in the pre-exposure population.

Our study has several limitations. Although we attempted to control for known confounders by using propensity score matching and multivariable adjustment, there is still a risk of the presence of uncontrolled, unknown confounding factors and bias due to the retrospective study design. Additionally, the small size of the cohort limited the number of variables that were included in the propensity score model; we have thus incorporated a list of the most crucial prognostic and demographic factors in our model (27). Second, there were numerous asymptomatic patients, and as a result the time from symptom onset to admission was not evaluated in our study. However, we obtained detailed information on the length of delay from the date of diagnosis to treatment initiation, which provides is an objective measure of treatment timing. Third, QT prolongation or retinopathy, the known adverse effects of hydroxychloroquine(28, 29) were not actively measured in our study; while this must be kept into account as in any patient receiving hydroxychloroquine, no serious adverse reactions including cardiac toxicity or retinopathy were observed in our study population. Fourth, the baseline characteristics of the HQ plus antibiotics treatment group and the conservative care group is heterogenous (**Table 1**). The conservative treatment group was composed of slightly milder patients. However, such differences in baseline has was balanced after propensity score matching. Lastly, few patients in HQ plus antibiotics group received Lop/R treatment (for 7.5% of total patients), and this may have caused some confounding effects on our measured clinical outcomes. However, duration of Lop/R use was accounted in our multivariate Cox analysis whose result did not show any difference from other adjusted methods.

## CONCLUSION

HQ with antibiotics was not associated with better clinical outcomes and did not reduce time to viral clearance, length of hospital stays, and duration of symptoms compared to conservative treatment in mild to moderate COVID-19 patients.

## Acknowledgements

No acknowledgements.

## Notes

**Declaration of Conflicting interest** The authors have no competing interests to disclose

### Competing Interest Statement

The authors have declared no competing interest.

### Author Declarations

The ethics committee of Pusan National University Yangsan Hospital approved this study and granted a waiver of informed consent from study participants.

